# Online trend estimation and detection of trend deviations in sub-sewershed time series of SARS-CoV-2 RNA measured in wastewater

**DOI:** 10.1101/2023.10.26.23297635

**Authors:** Katherine B. Ensor, Julia Schedler, Thomas Sun, Rebecca Schneider, Anthony Mulenga, Jingjing Wu, Lauren B. Stadler, Loren Hopkins

## Abstract

Wastewater surveillance has proven a key public health tool to understand a wide range of community health diseases and has proven to be especially critical to health departments throughout the SARS CoV-2 pandemic. The size of the population served by a wastewater treatment plant (WWTP) may limit the targeted insight about community disease dynamics. To investigate this concern, samples of wastewater were obtained at lift stations upstream of WWTPs within the sewer network. First, an online, semi-automatic time series model is fitted to the weekly measurements of WWTP samples to estimate the viral trend for the community and compared to the time series observations from the lift stations. Second, deviations from the WWTP trend are identified using an Exponentially Weighted Moving Average (EWMA) control chart. The analysis reveals that the lift stations display slightly different dynamics than the larger WWTP, highlighting the more granular insight gleaned from sampling sites which represent smaller populations. Discussion focuses on the use of our methods to support rapid public health decision-making based on additional, targeted samples in times of concern.

## 1. Introduction

Wastewater-based epidemiology (WBE) is a cost-effective and fast way to survey the transmission of disease in populations, and it has been widely applied for the monitoring of viral pathogens, including SARS-CoV-2 (Kisand et al., 2023; Olesen et al., 2021). Multiple studies have confirmed the correlation between SARS-CoV-2 wastewater monitoring data and COVID-19 clinical testing data (Hopkins et al., 2023b; Ciannella et al., 2023; Kasprzyk-Hordern et al., 2023; Kaya et al., 2022; Mao et al., 2020; Peccia et al., 2020; Wolken et al., 2023). Routine wastewater monitoring highlights SARS-CoV-2 dynamics and provides early notice of the emergence in the population served by the wastewater catchment areas (Cao and Francis, 2021; Karthikeyan et al., 2021; Kisand et al., 2023; Li et al., 2021; Vallejo et al., 2022; Zhao et al., 2022; Kirby et al., 2022). In short, wastewater epidemiology is an effective public health tool guiding meaningful interventions (Hopkins et al., 2023a).

Wastewater monitoring for diseases has predominantly involved the collection of samples from the influent of wastewater treatment plants (WWTPs). Monitoring centralized WWTPs is typically easier for municipalities to implement due to routine sampling that takes place at the treatment plants unrelated to WBE. Since these centralized WWTPs in large municipalities often serve large populations, it can be unclear where to take action when pathogens are detected.

Expanding spatial granularity of WBE surveillance can improve the accuracy and actionability of routine wastewater monitoring. Several prior studies focused on sampling sites upstream of the WWTP, such as lift stations, schools, universities, and hospitals (Castro-Gutierrez et al., 2022; Fielding-Miller et al., 2023; Gibas et al., 2021; Haak et al., 2022; Holm et al., 2022; Scott et al., 2021; Spurbeck et al., 2021). The WBE program for the City of Houston has routinely made use of sub-sewershed measurements (hou-wastewater epi.org/, 2023). Moreover, the nested sampling strategy was proposed to monitor the trend of SARS-CoV-2 in the catchment area and simultaneously identify and trace the community-level COVID-19 hotspots (Wolken et al., 2023; Wang et al., 2023; Yeager et al., 2021). The strategy highlighted the importance of sampling site selection, which can help develop a more sensitive and effective wastewater surveillance system for COVID-19 and other diseases.

Studies on the use of nested sampling strategy for wastewater monitoring remain limited, and more investigations are needed regarding the application of nested sampling on WBE to maximize the value of the data and improve the system efficiency (Acosta et al., 2022; D’Aoust et al., 2021; Holm et al., 2022; Wang et al., 2023; Yeager et al., 2021). D’Aoust et al. (2021) compared the wastewater SARS-CoV-2 surveillance results from two locations in a small, rural community with the community’s COVID-19 cases, and recommended sampling from lift stations, because they observed an overall higher and more stable SARS-CoV-2 load as compared to its downstream wastewater treatment lagoon. Holm et al. (2022) observed no statistical difference between SARS-CoV-2 viral concentrations in WWTPs and several upstream sampling sites, including lift stations and manholes, and recommended sampling sites that support larger populations. Acosta et al. (2022) observed a lower correlation between COVID-19 clinic data and SARS-CoV-2 RNA in wastewater samples from neighborhoods compared to samples from WWTPs, indicating that the current smaller wastewater monitoring system may induce more heterogeneous data that can reduce the sensitivity of the surveillance. Acosta et al. (2022); Haak et al. (2022) also indicated that issues associated with sample collection may decrease the stability and representativeness of the data collected at the community or neighborhood level.

Despite the potential community-wide benefits in wastewater monitoring at the sub-sewershed level, statistical analyses on this approach have been limited in their modelling assumptions and flexibility. Several analyses assume wastewater measurements over time to be independent (Acosta et al., 2022; D’Aoust et al., 2021; Holm et al., 2022) and compare differences between sewershed and sub-sewershed measurements using a simple group-level difference of means or pairwise correlations. Failure to account for time dependence may underestimate variability and inadequately capture the temporal dynamics of the wastewater measurements, where clear trends and patterns in the data exist. Within the wide literature of statistical models for SARS-CoV-2 wastewater longitudinal trends, most typically fall under the categories of autoregressive or regression-based models (Cao and Francis, 2021; Peccia et al., 2020; Jeng et al., 2023), SEIR models (McMahan et al., 2021; Fazli et al., 2021), or machine learning approaches such as the artificial neural network (ANN) of Li et al. (2021) or the time-series based machine learning approaches in Lai et al. (2023). Application of these models to the comparison of sub-sewershed time series has been sparse to nonexistent, and many may not be appropriate for detection of structural deviations away from downstream measurements.

Previously, cubic smoothing splines have been used by the City of Houston (Hopkins et al., 2023b; Stadler et al., 2020) to identify the sometimes rapidly changing signal in the noisy wastewater time series obtained through routine monitoring of the city wastewater. This spline model captures the trend and curvature of the trend in the nonlinear time series from the noisy wastewater measurements. However, smoothing splines cannot separate the inherent variability of the time series from the measurement process.

We conduct two statistical analyses that offer flexible, semi-automatic, and online estimation of wastewater dynamics between large WWTPs and sub-sewersheds. First, we model the wastewater measurements using a dynamic linear state-space time series model. This modelling framework allows for both online and retrospective estimation of wastewater trends accompanied with confidence bands to capture the precision of the estimates via the Kalman filter and smoother Shumway and Stoffer (2017). These estimates give broad insight into whether sub-sewershed time series give different information from large, centralized WWTP time series, as well as the ability to forecast future values. Second, we utilize tools from statistical process control (SPC) literature, namely exponentially weighted moving average (EWMA) control charts, to monitor whether even just one of the lift station measurements deviates significantly from the trend estimate for the larger WWTP, allowing for immediate online detection of community-specific spikes in SARS-CoV-2.

## 2. Methods

### 2.1. Data description

The City of Houston has 39 WWTPs, serving populations from approximately 500,000 to 600 individuals. Within the larger WWTPs, there are a number of lift station (LS) facilities where wastewater can be sampled and may serve to refine the geographic resolution provided by wastewater analysis. This work focuses on the largest WWTP that serves a population of roughly 551,150 people. Wastewater was sampled from May 24, 2021 through March 13, 2023 for four lift stations (see Figure 1) which are geographically contained within the large WWTP catchment area.

**Figure 1.**
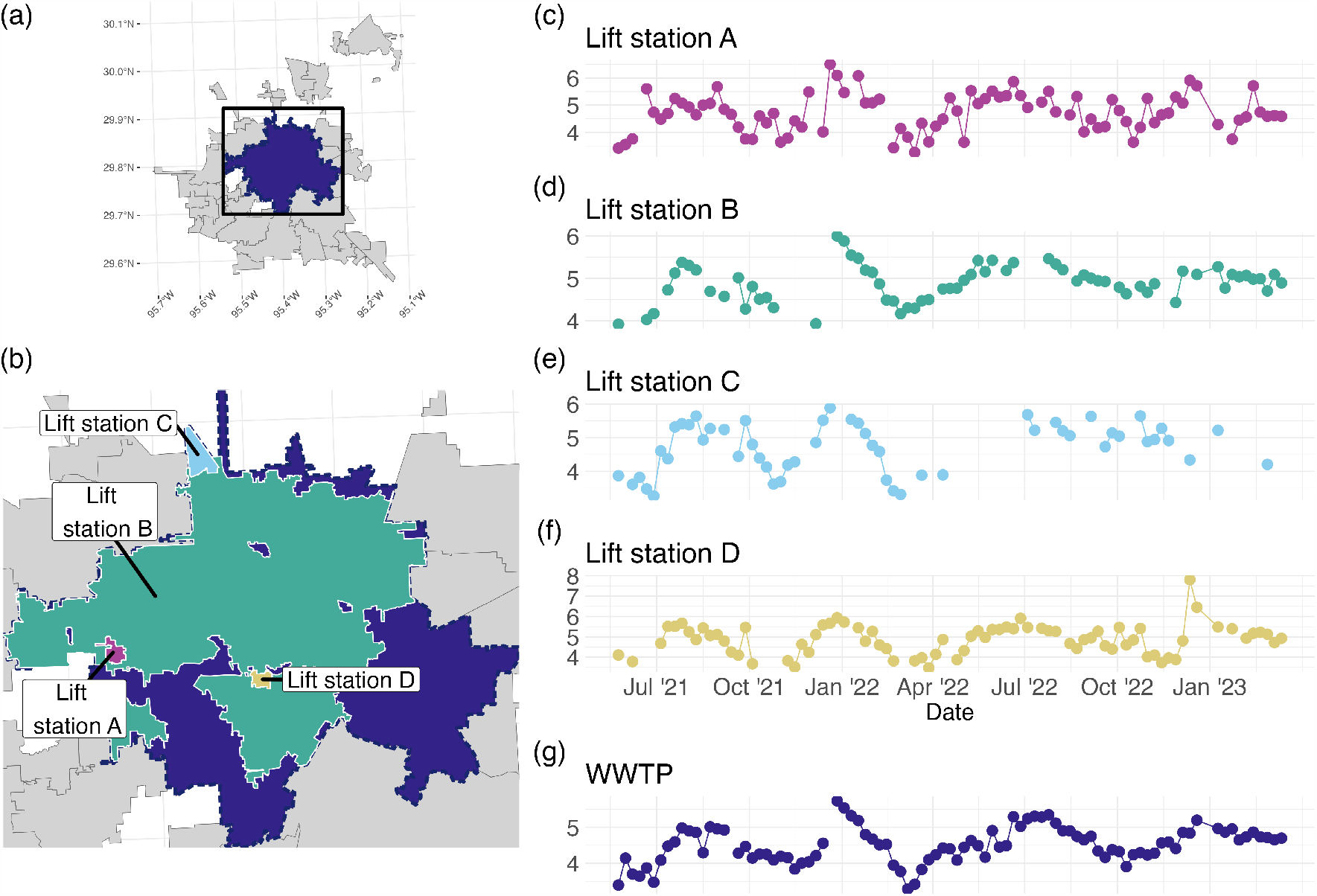
(a) The WWTP catchment areas for the City of Houston, with the WWTP of focus shaded. The box shows the extent of (b), the map showing the 4 lift stations considered in the analysis. (c)-(f) plot the time series of Log10 Copies/L for the WWTP and the 4 lift station facilities, referred to as Lift Station A-D.

Data on wastewater analysis results for the lift stations and the WWTP was updated on a weekly basis. For each weekly sample, we quantified SARS-CoV-2 N1 and N2 gene copies per liter of wastewater, as described previously (Hopkins et al., 2023b). We average the N1 and N2 concentrations to simplify our analysis and focus on the comparison between the WWTP and LS time series. All measurements taken in a given week were aligned to the corresponding Monday of that week. Wastewater viral concentration data was received in units of copies per liter and was subsequently log transformed in base 10 (log_10_). Any measurements below the level of detection (LOD) were labeled as missing values. Table 1 contains the names of the 5 series considered and summary statistics for each series. Figure 1 (a) and (b) are maps of the WWTP and LS catchments for each of the series, and Figure 1 (c)-(f) plot the time series of observed values for all 5 series on the log_10_ scale.

**Table 1:** Name, size of population, and summary statistics Log10 of average of replicate RNA N1 and N2 copies/L for each wastewater treatment plant (WWTP) or lift station (LS) considered. The study period spanned 93 weeks.

| Name | Population | Mean | St. Dev. | Min | Max | Missing |
| --- | --- | --- | --- | --- | --- | --- |
| WWTP | 551150 | 4.51 | 0.50 | 3.29 | 5.74 | 4 |
| Lift Station A | 2442 | 4.72 | 0.70 | 3.26 | 6.51 | 11 |
| Lift Station B | 373937 | 4.89 | 0.43 | 3.92 | 6.00 | 28 |
| Lift Station C | 4849 | 4.72 | 0.73 | 3.26 | 5.89 | 42 |
| Lift Station D | 1724 | 4.88 | 0.74 | 3.48 | 7.81 | 18 |

### 2.2. Hierarchical Time Series Model for Trend Estimation

When time series data are collected, the goal is often to estimate a trend, that is, whether the “typical values” are changing in time. For example, 1 (c)-(g) show the times series of viral concentration of SARS-CoV-2. Visually, it is clear that these values are changing in time, and even seem to exhibit similar behavior that may be predictable with a well-chosen model. Such a model should be able to separate out the “noise”, or observation/measurement error, in these observations from the “signal”, or trend. An additional constraint when modeling time series data is the presence of temporal correlation structure, i.e. the values are not independent, so models which assume independence can lead to misleading forecasts and/or conclusions about which variables are important in modeling a time series (Hyndman and Athanasopoulos, 2021). In summary, the desired model will separate sources of variability for both the trend and the observation as well as account for temporal correlation.

The state space modeling framework can accommodate both these needs. A state space model represents a time series in two levels: an unobserved trend which encodes temporal dependence structure and a noisy observed time series. In other words, it is a hierarchical model which is able to separate sources of variability as desired. In the time series literature, the levels of this model are called the state equation and the observation equation. Equations 1 and 2 display the state space model used for each series in this particular study:

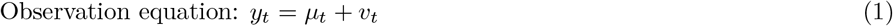

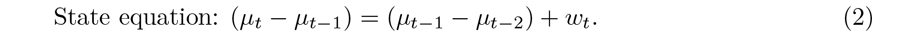

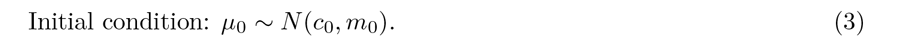

The error terms *v*_*t*_ and *w*_*t*_ are independent and normally distributed with mean zero, and variances denoted by 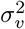 for the observation error and 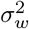 for the state error.

The observation model of Equation 1 represents the model fit to the concentration of SARS-CoV-2 RNA in wastewater measured by the lab. The observation model is the underlying state *μ*_*t*_ plus a variance term 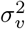 corresponding to the inherent measurement and sampling error. The state model in Equation 2 represents the true state of the viral trend derived from the measured concentration of SARS-CoV-2 RNA, for the sampled region. The noise term associated with the state equation, 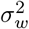, represents the natural variability in the viral concentration in the population as measured by wastewater.

Within this framework, the state variable serves as the core component of the model, characterizing the underlying system’s behavior and dynamics, in other words the trend of the virus concentration. Note the temporal structure encoded by Equation 2: the right hand side concerns the previous two time points, while the left hand side concerns the current and past time point. In particular, Equation 2 encompasses a statistical framework that employs the concept of first difference applied twice. The first difference operation captures the change in the state variable over successive time periods, and by applying this operation twice, we gain insights into the acceleration or curvature of the trend. That is, this choice of structure for the state equation is chosen to capture the temporal dependence of the SARS-CoV-2 RNA concentration. For additional details about the state-space modeling framework and its relations to smoothing splines, see Shumway and Stoffer (2017).

Once the structure of the model is chosen, the model can be fit to the data with three goals in mind: retrospective estimates of the trend using all available data, online estimates of the trend using only past data up to a given time point, and one-step-ahead forecasts of the next time point. In the time series literature, the retrospective and online estimates are referred to as smoothers and filters, respectively. We focus on retrospective and online estimates for this paper, but provide steps for obtaining the one-step-ahead forecasts in the supplemental materials.

To estimate the online and retrospective trends, four parameters are themselves estimated: the initial state mean and variance, the variance of the measurement and sampling error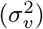, and the variance of the trend 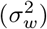. Estimates are obtained through maximum likelihood estimation which is computationally fast due to use of the Kalman Filter for updating linear Gaussian systems. For the online estimates, a rolling estimation structure is used, meaning the parameters are re-estimated with each new time point. Estimation is implemented using the KFAS package in R Helske (2017), which can easily handle missing data. Once an estimate of the model is obtained, a step to check that the model fits the data is required. For the present model, an autocorrelation plot of the model’s residuals can be checked for autocorrelation. If no correlation is present in the residuals, the model can be considered a good fit. Additional details of estimation and model fit checks as well as all code used for the analysis are available in the supplemental materials.

The inputs, outputs, and process of fitting the spline state space model of Equations 1 and 2 are summarized in Algorithm 1.

#### Algorithm 1: Variability-separating trend estimation

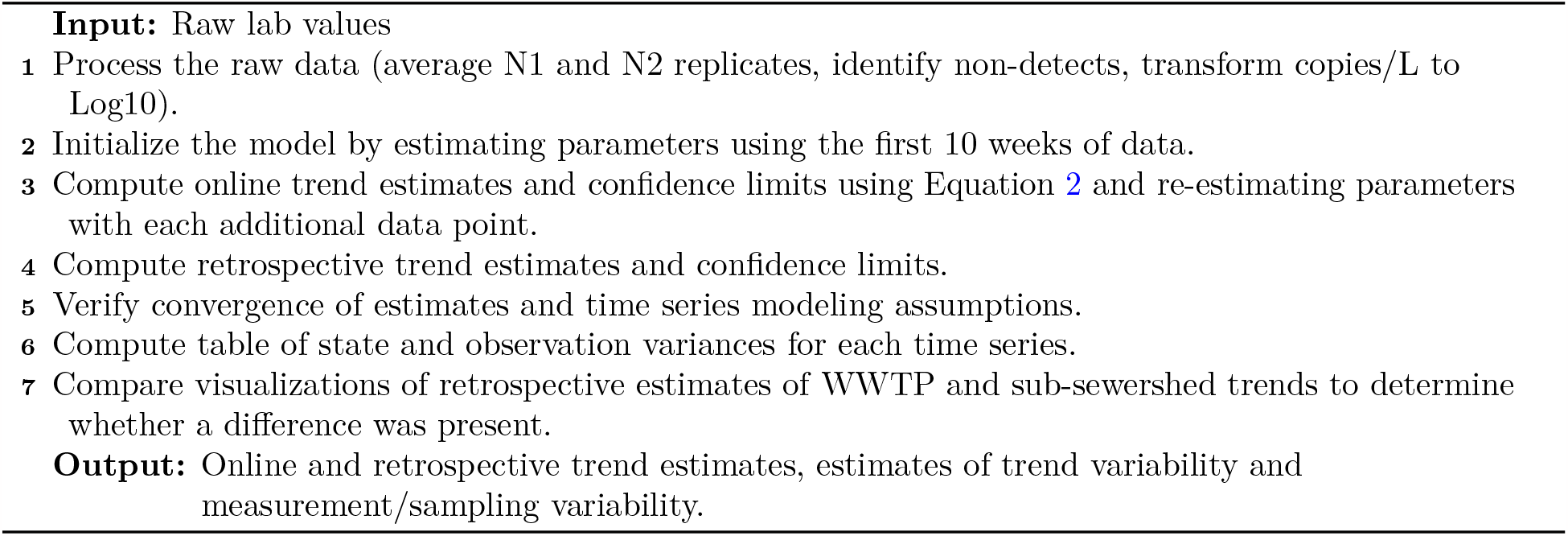

Visualizations of the retrospective and online estimates along with the data are provided in 2.

### 2.3. Detection of trend deviations

Recall the goal of determining whether sub-sewershed measurements give different information than the routinely monitored centralized WWTP measurements. Using all available data, the retrospective estimates from the model fit using Equations 1 and 2. These estimates, visualized in Figure 3, show some periods of separation, indicating that the sub-sewershed measurements do indeed give different information. However, if the goal is to extract actionable information from the data, the online estimates, which only use data up to the current time point, should be used. While the retrospective estimates show clear separation, the online estimates are noisier, so detecting when the sub-sewersheds may be deviating from the WWTP’s trend requires more than a visual comparison of the two series. In addition, sub-sewersheds may not be sampled frequently enough to support the model described in Section 2.2, so a method which can be used with at least one sub-sewershed observation is ideal.

**Figure 2.**
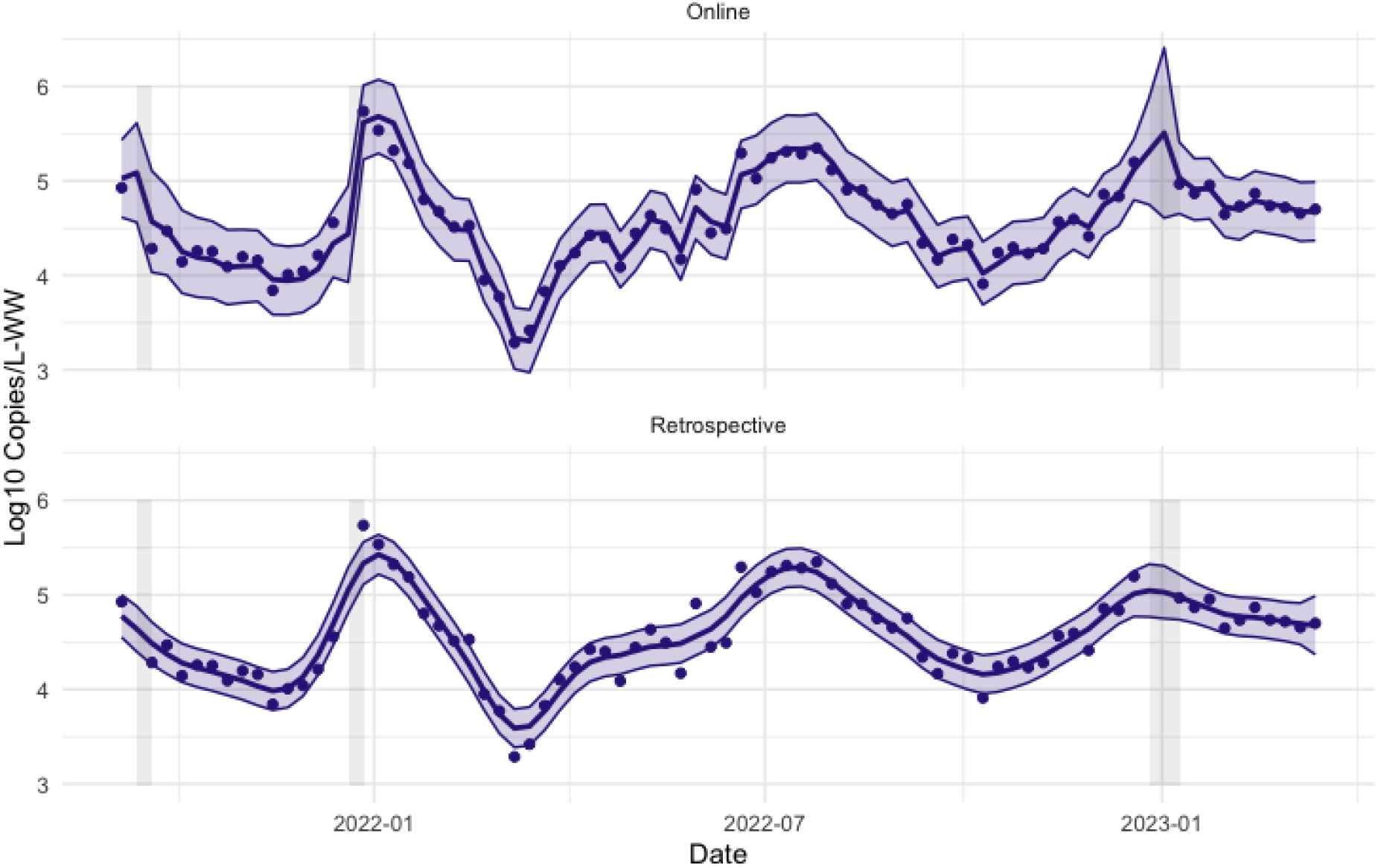
Retrospective and online estimates of the viral concentration trend with uncertainty quantification for the large WWTP. The vertical axis is log10 copies/liter. The shaded grey rectangles correspond to periods of missing data. Note that the online trend estimates are “noisier” and have wider uncertainty bands than the retrospective trend estimates.

**Figure 3.**
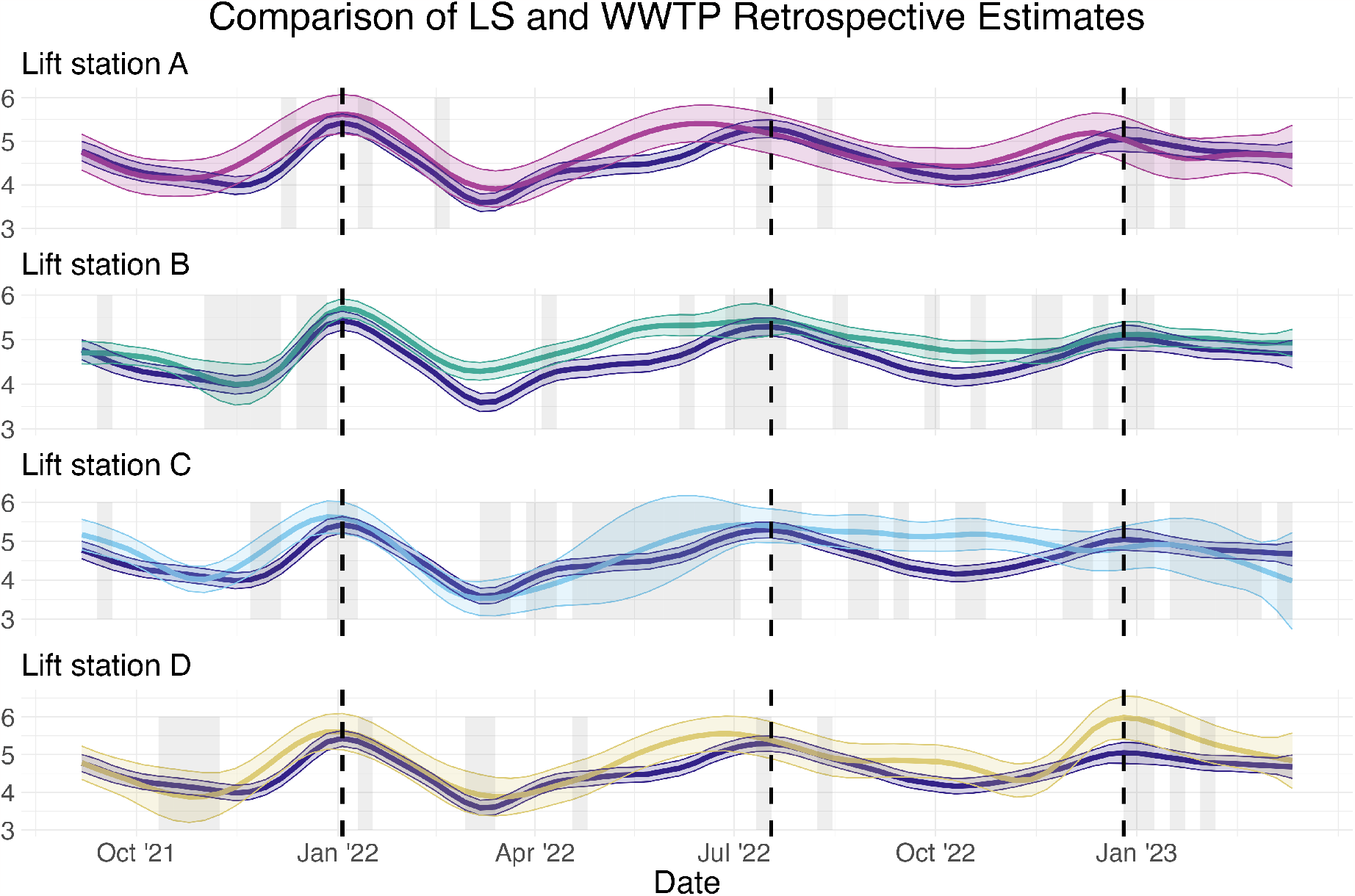
Retrospective estimates of the viral concentration trend with uncertainty quantification for the WWTP and each LS series, using all available information. The vertical axis is log10 copies/liter. The shaded grey rectangles correspond to periods of missing data and the dotted lines correspond to the peaks of three surges. Note that the time series model is still able to provide estimates of the trend during periods of missing data, though with greater uncertainty. Compared to Figure 1, the start date of this trend plot is later, since the first 10 weeks of data are used to initialize the model.

The statistical process control (SPC) literature provides a framework for iterative improvement of a decision-making process based on time series data. Some examples of the traditional applications of SPC include ensuring a given percentage of on-time deliveries to a client, speed and consistency of service quality in a bank, and loading passengers onto an airplane (Montgomery, 2009). In short, SPC provides a framework for identifying when a time series of interest is “out of control” so that steps can be taken to bring that series back “in control”. Although the ability to bring disease burden in a community back “in control” is limited in WBE compared to traditional applications, ideas from SPC can be borrowed to improve the actionability of the information contained in wastewater time series.

For this paper, the time series of interest is the difference between the sub-sewershed and the WWTP. If we simply subtract the observed values for each series, the resulting difference will contain the “noise”, or measurement and sampling error. Instead, we use the online estimate of the trend for the WWTP obtained from Equations 1 and 2, which can be assumed to be free of observation error. Since the online estimate of the trend requires 10 weeks of data to be initialized, we use the observed (unmodeled) value(s) from the sub-sewershed directly.

Formally, using the previous notation, the standardized difference at time point *t*, for lift station *i* = 1, …, 4 is given by:

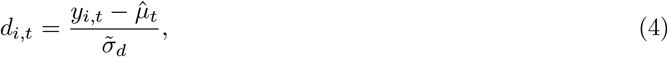

Where 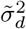 = Var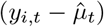. This variance is approximated by

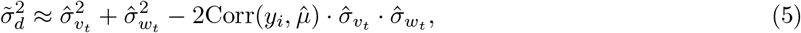

where Corr 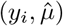 is the Pearson correlation coefficient between the WWTP estimated state time series and the observed copies/liter from the *i*^*th*^ lift station. If any of the sub-sewershed values *y*_*i,t*_ are missing, we replace these values with the online trend estimate for the WWTP, which will yield a value of 0.

If the sub-sewershed and the WWTP are “in control”, or gave equivalent information, then *d*_*i,t*_ would be normally distributed with mean 0, and there would be no autocorrelation in the series. To determine whether the sub-sewershed is “out of control”, or separating from the trend of the WWTP, a control chart can be constructed. Many types of control charts are available for different scenarios, for example, Shewhart (Shewhart, 1931) and CUSUM (Page, 1954) control charts. We choose an Exponentially Weighted Moving Average control chart (Roberts, 1959), which can detect small shifts in temporally correlated series such as our *d*_*i,t*_ and is appropriate for use with individual observations (Montgomery, 2009). The EWMA chart is based on the following series:

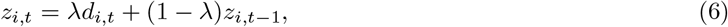

where *z*_*i,t*_ can be interpreted as a weighted average of all past values for series *i*, where the weighting is controlled by the value *λ*, for which we use the estimate of the lag 1 autocorrelation of *d*_*i,t*_. In the case of a missing sub-sewershed value, the aforementioned replacement with the WWTP online estimate allows for the exponential weighting of past values to continue under the assumption of no separation. The EWMA charts are visualized for each of the 4 lift stations compared to the WWTP in Figure 4.

**Figure 4.**
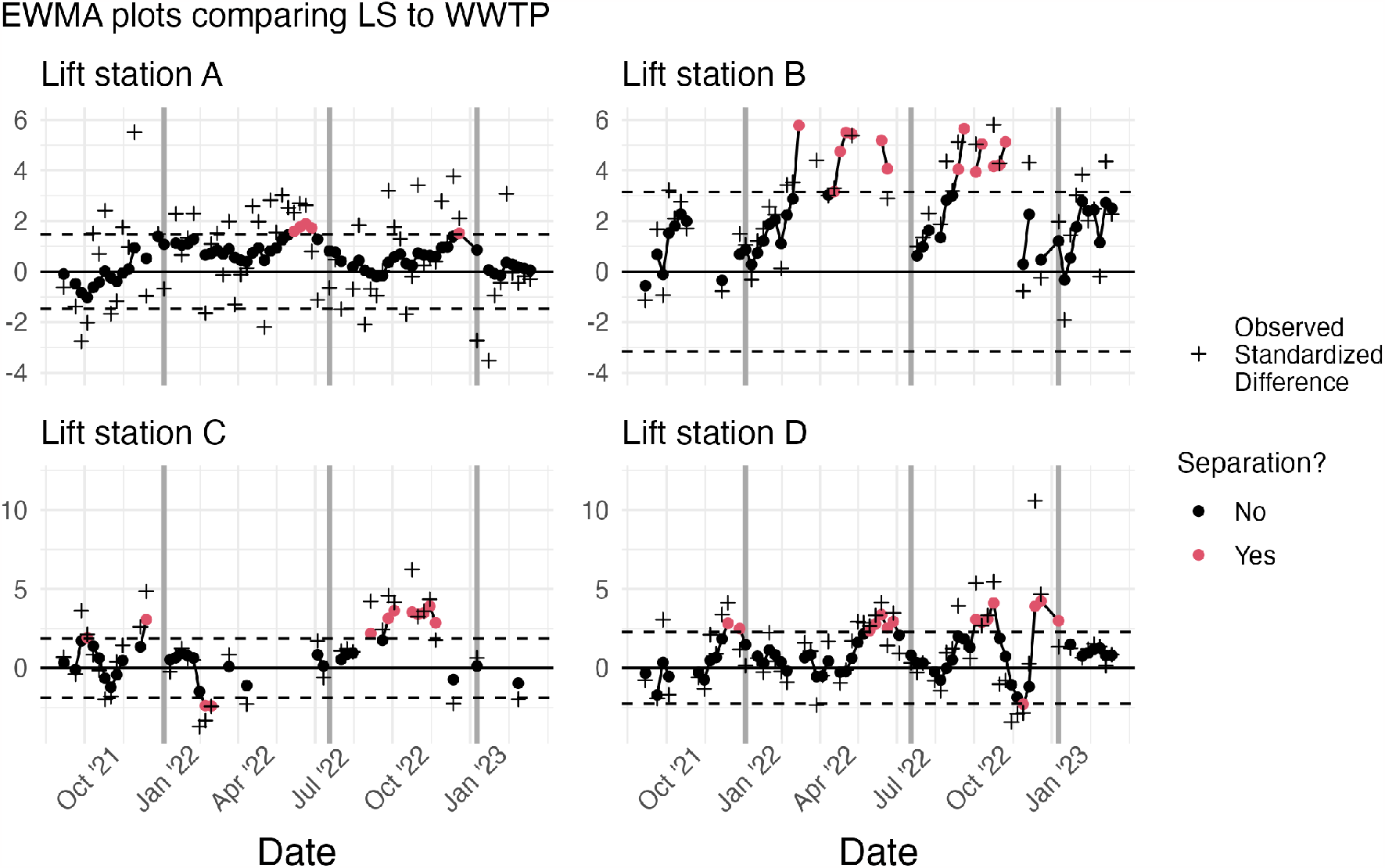
The EWMA chart for the observed values at each lift station compared to the WWTP online estimate. The solid dots represent the exponentially weighted standardized difference while the plus signs represent the actual standardized difference. Observations which correspond to a structural break, or exponentially weighted values beyond the dotted control limits, are colored red. The dark grey vertical lines are the approximate dates of the peaks of different surges.

The dots on Figure 4 represent the values of *z*_*t*_. The dotted lines are the upper and lower confidence limits. When *z*_*t*_ exceeds one of these confidence limits, the point is colored red, and the sub-sewershed can be considered “out of control”, in other words, the sub-sewershed time series is separating from the WWTP time series which gives different information. The direction of the separation can also be determined by examining whether the point exceeds the upper limit, indicating the viral concentration is higher for the sub-sewershed, or the lower limit, indicating the sub-sewershed is lower.

We summarize the creation of the EWMA chart in Algorithm 2. For additional discussion of EWMA charts and examples of their use with correlated data, see Hunter (1986); Lucas and Saccucci (1990); Supharakonsakun et al. (2020).

#### Algorithm 2: Detecting deviation of sub-sewershed measurement from centralized WWTP trend estimate

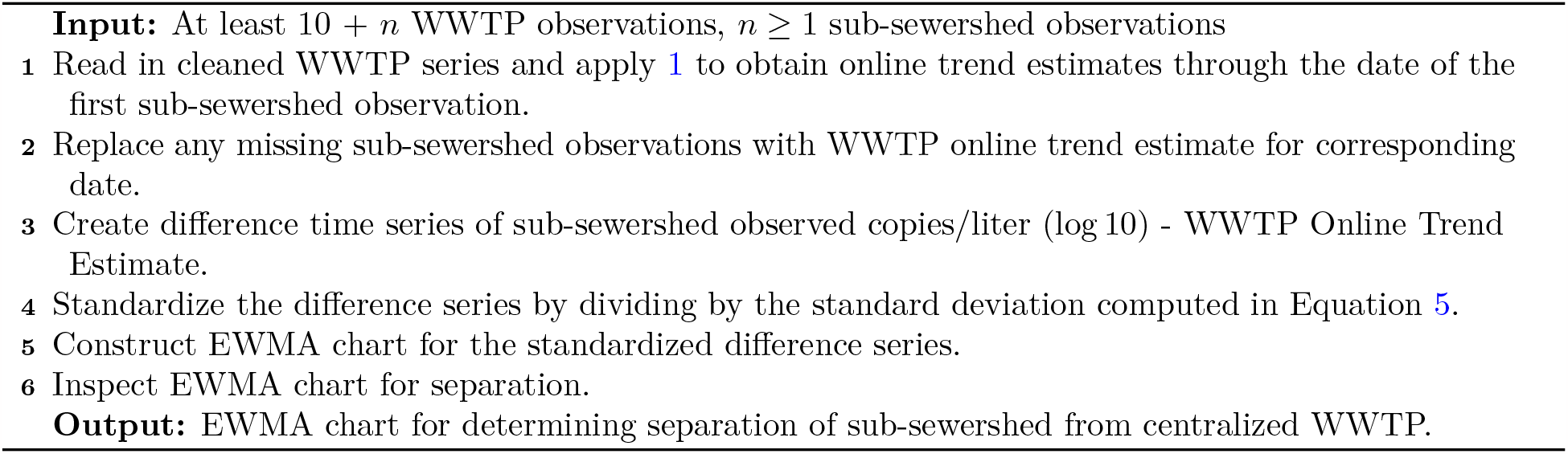

## Data Availability

Given the small populations associated with some of the lift stations, real data will be made available on a case-by-case basis by contacting the corresponding author and subsequent approval by Houston Health Department. Synthetic wastewater surveillance data which preserves the statistical properties of the real data are available along with code on a GitHub repository.

## Code Availability

All code used to fit the models described in this paper are available in a public GitHub repository. All code is written in the R language (R Core Team, 2023).

## 3. Results

### 3.1. Trend Estimation

The retrospective estimates depicted in Figure 3 indicates three peaks in the estimated population viral dynamics for the population served by the WWTP, with maximums that occur on January 3 and July 18, 2022, and January 9, 2023. We will refer to these peaks as PK1, PK2 and PK3, respectively. The retrospective review illustrates there are instances where the lift stations provided early information with respect to increasing or decreasing viral trends in the population measured. Again, these features are highlighted in Figure 4. A separation in the confidence intervals for each series indicates a statistically significant difference in the estimated trend for the respective series, namely the trend estimated for the WWTP and each of the lift stations.

We also see in the retrospective review that three of the four lift stations, namely Lift station A, Lift station C, and Lift station D, exhibit a comparable trend as that estimated from the WWTP, with a few deviations (see Figure 3). Lift station A indicates an early signal leading up to PK1. Lift station C separates from the WWTP trend by remaining at high levels between PK1 and PK2. It is important to note that there are several missing values in the Lift station A series during this time, as highlighted by the light grey bars in Figure 3. However, the estimated level for Lift station C does not show an increase in uncertainty due to the fact that the observed levels during this time were relatively consistent (see Figure 1). Lift station D registers a statistically higher trend in PK3. Lift station B was unique amongst the four lift stations, in that its estimated trend separated from the estimated WWTP trend following PK1 and remained higher until PK2. Lift station B also failed to drop as low between PK2 and PK3. However, the trend estimates for the WWTP and Lift station B were not significantly different in PK3. It is also worth noting that Lift station A, Lift station C, and Lift station D exhibited more variation in their trend estimates as evidenced by the width of the respective confidence intervals, than that of the WWTP.

Recall that the hierarchical trend estimation framework can separate variability associated with the trend from the “noise”, or lab and sampling error. These results are summarized in Table 2. We see from this table that the measurement and sampling variation are highest for Lift station A and Lift station D, and also elevated for Lift station C. Since we expect the lab variability to be approximately constant across all measurements, the extra variation is most likely due to the lift station wastewater containing highly variable levels of SARS-CoV-2 due to the small population that it serves. Based on this observation, the sampling variation is approximately equivalent for Lift station B as it is for the WWTP. The state dynamics for each location exhibit similar variability, with slightly elevated variation for Lift station D.

**Table 2:** Estimates of inherent variability, *σ*_*w*_ (state) and measurement variability *σ*_*v*_ (observation, lab and sampling variability) for each series.

| Name | Sampling and Lab<br>Variability | Trend<br>Variability | Population |
| --- | --- | --- | --- |
| WWTP | 0.0372 | 0.0130 | 551150 |
| Lift station A | 0.2798 | 0.0105 | 2442 |
| Lift station B | 0.0350 | 0.0130 | 373937 |
| Lift station C | 0.1374 | 0.0134 | 4849 |
| Lift station D | 0.2810 | 0.0175 | 1724 |

Again, the retrospective review provides the best understanding of the dynamic population trend in virus levels for location as well as insight into the lab and sampling variability over the entire study period. However, this retrospective review is not useful in real time as it requires knowledge of the full time series, i.e. the future, to implement.

### 3.2. Trend Deviations

The results of Algorithm 2 applied to each lift station and the WWTP are graphically introduced in Figure 4. The information is consistent with the retrospective study in that Lift station A, Lift station C and Lift station D, all demonstrate minor perturbations from the trend estimated for the WWTP. Further, Lift station B clearly demonstrates a strong and consistent deviation from the WWTP estimated trend, between PK1 and PK2, and then again between PK2 and PK3. In Figure 4 we also include the observed standardized difference between the two measurements. You will note that the differences may be large, but they are not statistically significant based on the EWMA control chart. The control chart is used to identify a level shift in the trend, and not specific outlying events. Based on the EWMA control chart, statistically significant level shifts occurred at the red highlighted temporal locations.

## 4. Discussion

The objective of this paper is to highlight the additional information gleaned from samples taken within the sewer network and upstream of WWTPs, namely lift stations. We bring forward a hierarchical time series approach to capture the dynamic trend in population viral dynamics from each wastewater series. The state-space time series model is simple to implement both in a retrospective, and real-time mode, and naturally adapts to the nonlinear dynamics in the population viral trend. The EWMA control charts provide a framework for identifying when sub-sewershed measurements deviate from measurements in a larger, centralized WWTP. Note that these deviations could be due to any number of factors, for example, number of infections, dilution, degradation, inhibitors, etc.

For our system the only lift station within the large wastewater catchment area of the WTTP whose trend consistently deviated from that of the WWTP was Lift station B. Further, the measurement and sampling uncertainty for Lift station B was on par with that of the WWTP. This lift station serves 373,937 people whereas the WWTP serves 551,150 people. In other words, Lift station B serves 68% of the people in the large catchment area. Regular monitoring of Lift station B in addition to the WWTP is warranted based on this study. The Lift station B state estimate of viral load and its uncertainty, indicates that for the 68% of the population served by Lift station B, the viral load did not decrease as substantially as that of the WWTP between PK1 and PK2, and also between PK2 and PK3.

For the lift stations serving smaller populations, namely Lift station A, Lift station C and Lift station D, we see evidence of early signals through each COVID-19 peak. However, the measurement and sampling uncertainty with these smaller lift stations was substantially higher. Although routine monitoring may be prohibitively expensive, monitoring through times of high concern to public health may be warranted.

A side result of our modeling approach is the opportunity to separate the variation in the trend of the viral load from the measurement and sampling variation. In this comparison, and assuming a consistent measurement or lab variability across all samples, we find that the sampling variation for the smaller lift stations is much greater than that for the WWTP and the large Lift station B. If regular sampling at smaller lift stations, where flow may be irregular, is required, sampling strategies may need to be reviewed.

## Data Availability

Given the small populations associated with some of the lift stations, real data will be made available on a case-by-case basis by contacting the corresponding author and subsequent approval by Houston Health Department. Synthetic wastewater surveillance data which preserves the statistical properties of the real data are available along with code on a GitHub repository (https://github.com/hou-wastewater-epi-org/online_trend_estimation).

## Acknowledgements

The authors disclosed receipt of the following financial support for the research, authorship, and/or publication of this article: This work was supported by the CDC Foundation (project no. 1085.46) and the Centers for Disease Control and Prevention (ELC-ED grant no. 6NU50CK000557-01-05 and ELC-CORE grant no. NU50CK000557). The work was also supported by the Rockefeller Foundation.

